# The Ontario Neurodegenerative Disease Research Initiative

**DOI:** 10.1101/2020.07.30.20165456

**Authors:** Kelly M. Sunderland, Derek Beaton, Stephen R. Arnott, Peter Kleinstiver, Donna Kwan, Jane M. Lawrence-Dewar, Joel Ramirez, Brian Tan, Robert Bartha, Sandra E. Black, Michael Borrie, Donald Brien, Leanne K. Casaubon, Brian C. Coe, Benjamin Cornish, Allison A. Dilliott, Dar Dowlatshahi, Elizabeth Finger, Corinne Fischer, Andrew Frank, Julia Fraser, Morris Freedman, Barry Greenberg, David A. Grimes, Ayman Hassan, Wendy Hatch, Robert A. Hegele, Christopher Hudson, Mandar Jog, Sanjeev Kumar, Anthony Lang, Brian Levine, Wendy Lou, Jennifer Mandzia, Connie Marras, William McIlroy, Manuel Montero-Odasso, David G. Munoz, Douglas P. Munoz, Joseph B. Orange, David S. Park, Stephen H. Pasternak, Frederico Pieruccini- Faria, Tarek K. Rajji, Angela C. Roberts, John F. Robinson, Ekaterina Rogaeva, Demetrios J. Sahlas, Gustavo Saposnik, Christopher J.M. Scott, Dallas Seitz, Christen Shoesmith, Thomas D.L. Steeves, Michael J. Strong, Stephen C. Strother, Richard H. Swartz, Sean Symons, David F. Tang-Wai, Maria Carmela Tartaglia, Angela K. Troyer, John Turnbull, Lorne Zinman, Paula M. McLaughlin, Mario Masellis, Malcolm A. Binns, on behalf of ONDRI Investigators

## Abstract

**Objective:** In individuals over the age of 65, concomitant neurodegenerative pathologies contribute to cognitive and/or motor decline and can be aggravated by cerebrovascular disease, but our understanding of how these pathologies synergize to produce the decline represents an important knowledge gap. The Ontario Neurodegenerative Disease Research Initiative (ONDRI), a multi-site, longitudinal, observational cohort study, recruited participants across multiple prevalent neurodegenerative diseases and cerebrovascular disease, collecting a wide array of data and thus allowing for deep investigation into common and unique phenotypes. This paper describes baseline features of the ONDRI cohort, understanding of which is essential when conducting analyses or interpreting results.

**Methods:** Five disease cohorts were recruited: Alzheimer’s disease/amnestic mild cognitive impairment (AD/MCI), amyotrophic lateral sclerosis (ALS), frontotemporal dementia (FTD), Parkinson’s disease (PD), and cerebrovascular disease (CVD). Assessment platforms included clinical, neuropsychology, eye tracking, gait and balance, neuroimaging, retinal imaging, genomics, and pathology. We describe recruitment, data collection, and data curation protocols, and provide a summary of ONDRI baseline characteristics.

**Results:** 520 participants were enrolled. Most participants were in the early stages of disease progression. Participants had a median age of 69 years, a median Montreal Cognitive Assessment score of 25, a median percent of independence of 100 for basic activities of daily living, and a median of 93 for instrumental activities. Variation between disease cohorts existed for age, level of cognition, and geographic location.

**Conclusion:** ONDRI data will enable exploration into unique and shared pathological mechanisms contributing to cognitive and motor decline across the spectrum of neurodegenerative diseases.

## Introduction

With heterogeneous symptoms that interfere with daily functioning, including cognitive impairment and/or the deterioration of motor abilities, neurodegenerative diseases take a substantial toll on the quality of life for patients and their families, and results in a myriad of burdens across social, economic, and healthcare systems^1^.

Several large studies examine specific neurodegenerative diseases^2–5^. These include cohort studies such as the Alzheimer’s Disease Neuroimaging Initiative (ADNI) and the Parkinson Progression Marker Initiative (PPMI), which have advanced our understanding of prototypical Alzheimer’s disease (AD) and Parkinson’s disease (PD), respectively. However, there is growing awareness that pathological mechanisms may overlap among neurodegenerative diseases^6-9^, especially in relation to the development of cognitive and motor decline. Furthermore, neurodegeneration can be exacerbated by cerebrovascular disease (CVD)^10^. The Ontario Neurodegenerative Disease Research Initiative (ONDRI) is a longitudinal, multi-site, observational cohort study undertaken to enable the exploration and extend our knowledge of similarities and differences within and among neurodegenerative conditions, their relationship with cerebrovascular disease, and potential synergistic mechanisms yielding cognitive and motor decline^11^. Participants were recruited into five disease cohorts: (1) AD and amnestic mild cognitive impairment (MCI), (2) amyotrophic lateral sclerosis (ALS), (3) frontotemporal dementia spectrum disorders (FTD), (4) PD, and (5) CVD, and were assessed annually with a rigorous set of measurement tasks across seven diverse assessment platforms. Herein, we describe the recruitment and characteristics of the ONDRI cohort at baseline allowing for more nuanced analyses of these data and appropriate interpretation of subsequent publications.

## Methods

### Eligibility and recruitment

Participants were recruited through tertiary clinics at fourteen academic health science centres in six cities across Ontario, Canada. Figure 1 lists general and platform-specific inclusion and exclusion criteria; additional details are elsewhere^11^. Eligible participants were previously diagnosed with one of the ONDRI-focused diseases and met consensus diagnostic criteria that was contemporary at the time of enrolment: AD^12^, including both amnestic and non-amnestic presentations; MCI with single- or multi-domain amnestic involvement^13^; possible, probable, or definite ALS, based on revised El Escorial criteria^14^; FTD spectrum disorders, including behavioural variant FTD^15^, corticobasal syndrome^16^, progressive supranuclear palsy^17^, and progressive primary aphasia, including agrammatic/non-fluent and semantic variants^18^ but not the logopenic variant; idiopathic PD with or without cognitive impairment^19^; or CVD with or without cognitive impairment. Participants eligible for the CVD cohort experienced a mild to moderate ischemic stroke/transient ischemic attack (TIA) (individuals with large cortical strokes were excluded) confirmed by clinical imaging (CT and/or MRI at least three months prior to enrolment.

**Figure 1:**
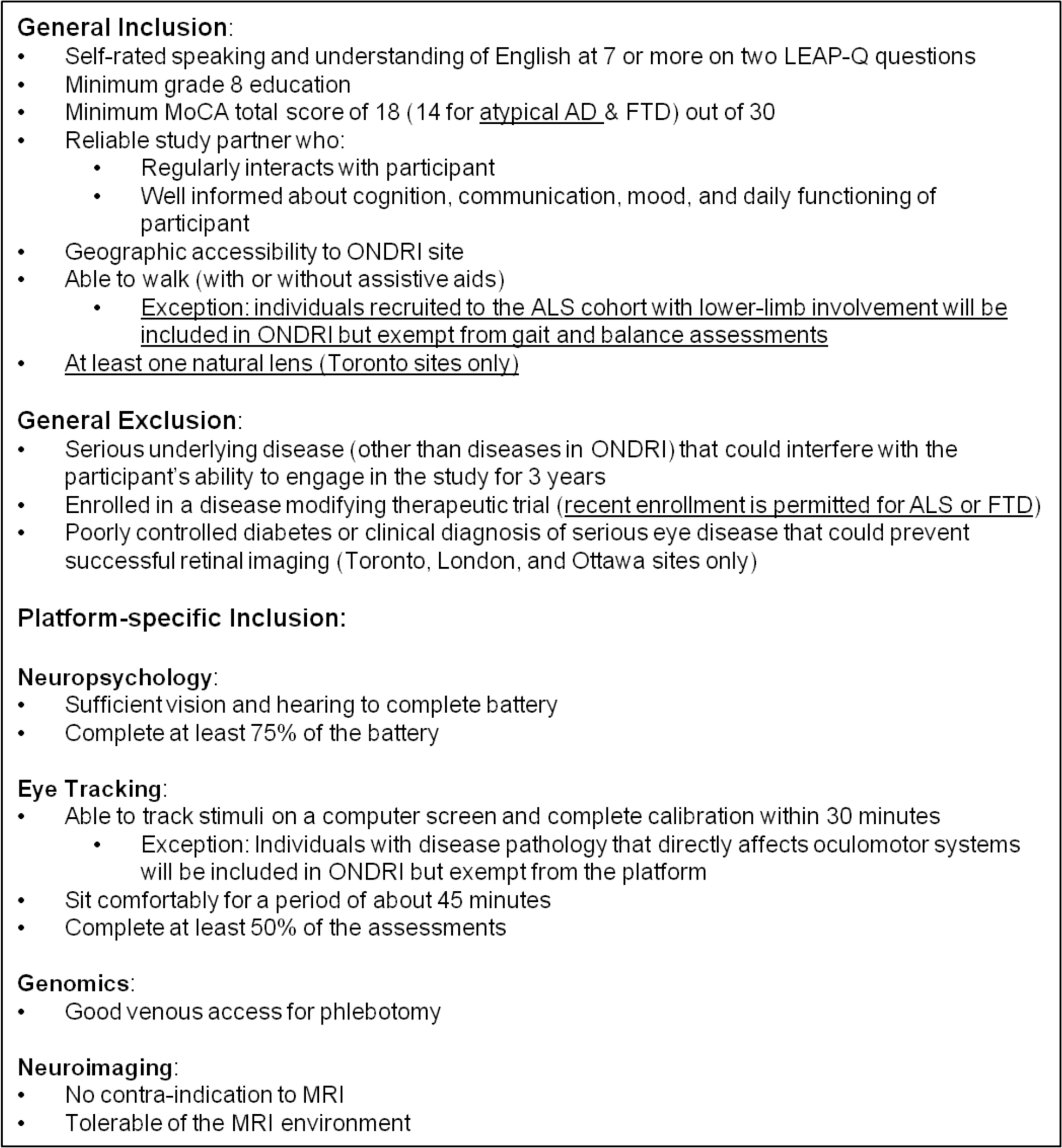
General and platform-specific inclusion and exclusion criteria. Any changes from the previous publication^11^ are underlined. LEAP-Q = Language Experience and Proficiency Questionnaire; MoCA = Montreal Cognitive Assessment; AD = Alzheimer’s disease; FTD = frontotemporal dementia spectrum disorder; ALS= amyotrophic lateral sclerosis.

MRI scans, collected according to a standardized Canadian protocol^20^ as part of the neuroimaging assessment, for all participants recruited to the AD/MCI cohort were assessed before enrolment by a research neuroradiologist to confirm absence of significant pathology that suggested a non-Alzheimer’s related cause of cognitive impairment^21^.

The study aimed to enrol individuals with clinical characteristics that would maximize the ability to observe disease progression and permit the identification of differences and/or similarities between the various trajectories of aging with neurodegenerative disease, especially with respect to cognition. As a result, disease-specific inclusion criteria targeted individuals in the earlier stages of disease, and clinicians recruiting for the CVD or PD cohorts were encouraged to enrol both cognitively intact participants and participants with some evidence of cognitive impairment/dementia, as defined by a cut-off of 26 on the Montreal Cognitive Assessment (MoCA)^22^. Further, clinicians recruiting to the PD cohort were encouraged to enrol more participants 65 years of age or older (a proportion of two-thirds) than those younger than 65 years as changes in cognition were more likely to be observed in an older population.

A healthy control cohort with overlapping assessments to ONDRI has also been collected from the Brain-Eye Amyloid Memory study^23^. Baseline characteristics of this control cohort will be reported in a separate paper.

### Assessment platforms

Participants completed each assessment platform annually. To account for the more rapid motoric and respiratory decline of individuals with ALS, participants in that cohort completed clinical, neuropsychology, and eye tracking assessments at 6-month intervals. Additional information pertaining to the assessment platforms is provided elsewhere^11,24-31^.

To ensure meaningful comparisons, participants were required to be able to undertake all assessments, except for retinal imaging, which was not available to participants enrolled in Kingston, Hamilton, or Thunder Bay, and those noted in Figure 1.

Study coordinators aimed to administer all baseline assessments within eight weeks of the participant providing written consent to optimize cross-platform comparisons and to minimize potential symptom-progression confounds (i.e., decline in status over time)^32^.

### Data management and quality control

All data are stored in Brain-CODE^33^: a secure, centralized neuroinformatics system designed for the collection, storage, federation, sharing, and analysis of different types of data using a diverse set of electronic data capture tools including XNAT, RedCap, and LabKey. Rigorous, assessment-specific data pre-processing and quality control procedures were developed and executed to ensure data were of the highest-possible fidelity^26–31^. This included investigation of any missing data and the use of detailed, non-numeric missing codes to indicate broad categories for missing values where they could be determined. All data packages were formatted to meet structure specifications, facilitating cross-platform merging and third party sharing. Subsequently, an ONDRI-wide data quality evaluation process was performed for each dataset. This procedure included multivariate outlier detection to identify data patterns and detect incongruous observations, consequently guiding toward areas of potential error^34,35^.

### Analysis of participant characteristics

Demographic and study partner data, as well as cross-disease and disease-specific clinical variables, were evaluated to describe the ONDRI sample at baseline. Demographic variables include age, sex, highest education level achieved, ethnicity, marital status, living arrangement, and region of recruitment. Study partner variables include relationship to participant, age, sex, education, whether they live with the participant, and time spent with participant. Disease-specific measures include age at first symptoms and duration of disease at time of recruitment, MoCA total score, Lawton-Brody activities of daily living (ADL) scales, neuropsychiatric inventory questionnaire (NPI-Q), Movement Disorder Society revised - Unified Parkinson’s Disease Rating Scale (S-UPDRS), modified Rankin scale (mRS), World Health Organization quality of life abbreviated scale (WHOQOL-BREF), revised ALS functional rating scale (ALSFRS-R) for the ALS cohort, and Hoehn & Yahr rating for the PD cohort.

Since this is an observational study, important clinical and demographic measures are not independent. In order to describe the multivariate relationships between these measures, we performed a Multiple Correspondence Analysis (MCA)^36^. For missing values, a sensitivity analysis was performed to assess the change in results given a range of plausible values before selecting the most appropriate with regression imputation. Analyses were performed with R version 3.4.4, including the ExPosition (v2.8.19)^37^ and ggplot2 (v3.1.0)^38^ packages.

We summarized frequency and type of missing data and time windows of baseline assessment completion to facilitate interpretation of future analyses: the frequency of participants missing all data within a platform and the reasons thereof influence cross-platform comparisons, while the temporal distance of data across platforms induces potential symptom-progression confound. Data collection time windows were calculated based on the date the participant provided written consent and the dates on which each assessment platform was completed, the date the demographic variables were captured for the clinical platform, and the date the majority of the tasks were completed for the neuropsychology platform.

### Standard protocol approvals, registrations, and patient consents

Research ethics committees at all participating recruitment sites approved the ONDRI research protocol. All participants and study partners provided written and informed consent prior to contribution and in accordance with the Declaration of Helsinki.

### Data availability

All ONDRI data, including but not limited to data used in this article, will be available to the scientific community through the Ontario Brain Institute later this year. Please see the Ontario Brain Institute website^39^ for more information.

## Results

### Recruitment

Participants were recruited between July 2014 and March 2017. Some inclusion and exclusion criteria were amended during the course of the study resulting in a broadening of the field of eligible participants.

In total, 630 individuals provided written informed consent to participate in ONDRI and were screened. Ultimately, 25 (4%) withdrew consent during screening and 94 (18%) did not meet inclusion criteria (Figure 2). However, 9 (1%) met inclusion criteria for a different cohort and were subsequently enrolled there.

**Figure 2:**
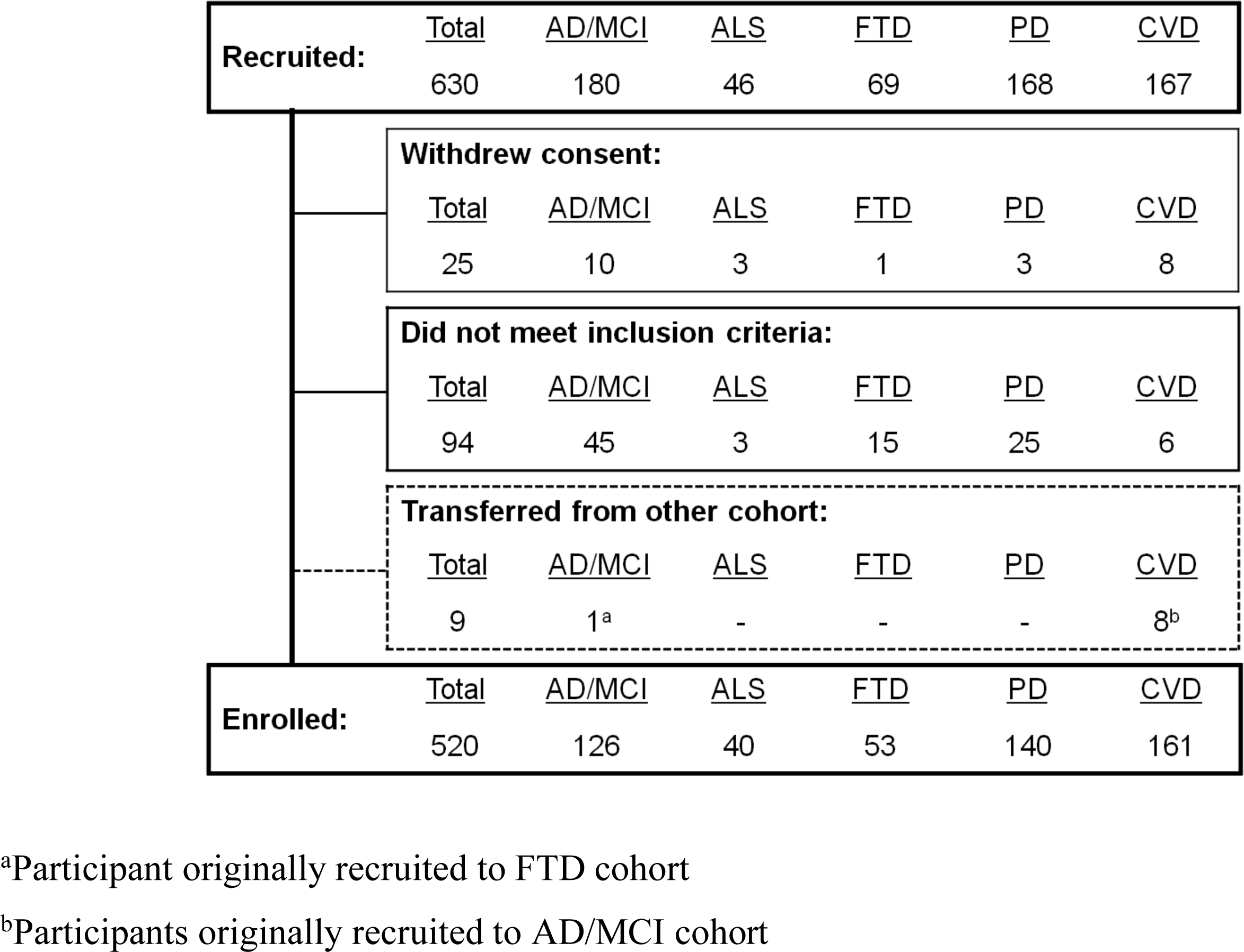
Consort diagram for individuals recruited to ONDRI. AD/MCI = Alzheimer’s disease and amnestic mild cognitive impairment; ALS = amyotrophic lateral sclerosis; FTD = frontotemporal dementia spectrum disorder; PD = Parkinson’s disease; CVD = cerebrovascular disease. ^a^Participant originally recruited to FTD cohort ^b^Participants originally recruited to AD/MCI cohort

Table 1 summarizes the inclusion criteria not met by 94 of the recruited individuals. Nine individuals were excluded from the disease cohort to which they were recruited based on MRI results challenging disease-specific requirements: 8 participants were excluded from the AD/MCI cohort (2 with Alzheimer’s disease and 6 with mild cognitive impairment) due to sufficient vascular pathology was identified, motivating their transfer to the CVD cohort^21^; and 1 participant was transferred from the FTD cohort (behavioural variant) to the AD/MCI cohort after prominent hippocampal atrophy instead of frontotemporal involvement was identified, with further historical clarification showing evidence of memory decline as the presenting symptom, in addition to behavioural dysregulation.

**Table 1:** Number (proportion) of individuals that did not meet inclusion criteria for criterion. MoCA = Montreal Cognitive Assessment; AD/MCI = Alzheimer’s disease and amnestic mild cognitive impairment; ALS = amyotrophic lateral sclerosis; FTD = frontotemporal dementia spectrum disorder; PD = Parkinson’s disease; CVD = cerebrovascular disease.

| **Criterion** | **Total** | **AD/MCI** | **ALS** | **FTD** | **PD** | **CVD** |
| --- | --- | --- | --- | --- | --- | --- |
| **Total** | 94 | 45 | 3 | 15 | 25 | 6 |
| **General Criteria** | 44 (47%) | 18 (40%) | 0 (0%) | 12 (80%) | 13 (52%) | 1 (17%) |
| • Low MoCA total score | 17 | 9 | 0 | 5 | 3 | 0 |
| • No study partner | 8 | 1 | 0 | 3 | 3 | 1 |
| • Underlying disease or condition | 5 | 3 | 0 | 0 | 2 | 0 |
| • Geographic accessibility | 3 | 0 | 0 | 2 | 1 | 0 |
| • Other | 11 | 5 | 0 | 2 | 4 | 0 |
| **Disease Cohort Criteria** | 21 (22%) | 16 (36%) | 0 (0%) | 1 (7%) | 4 (16%) | 0 (0%) |
| **Assessment Platform Criteria** | 29 (31%) | 11 (24%) | 3 (100%) | 2 (13%) | 8 (32%) | 5 (83%) |
| • Neuropsychology | 2 | 1 | 1 | 0 | 0 | 0 |
| • Eye tracking | 6 | 2 | 0 | 0 | 4 | 0 |
| • Neuroimaging | 21 | 8 | 2 | 2 | 4 | 5 |
| ○ MRI contraindication | 11 | 4 | 0 | 2 | 3 | 2 |
| ○ Unable to tolerate MRI | 7 | 2 | 2 | 0 | 0 | 3 |
| ○ Other | 3 | 2 | 0 | 0 | 1 | 0 |

### Participant and study partner characteristics

In total, 520 participants were enrolled in ONDRI across five cohorts: AD/MCI=126, ALS=40, FTD=53, PD=140, and CVD=161. Table 2 summarizes demographic variables for participants and Table 3 describes study partners for each disease cohort.

**Table 2:**
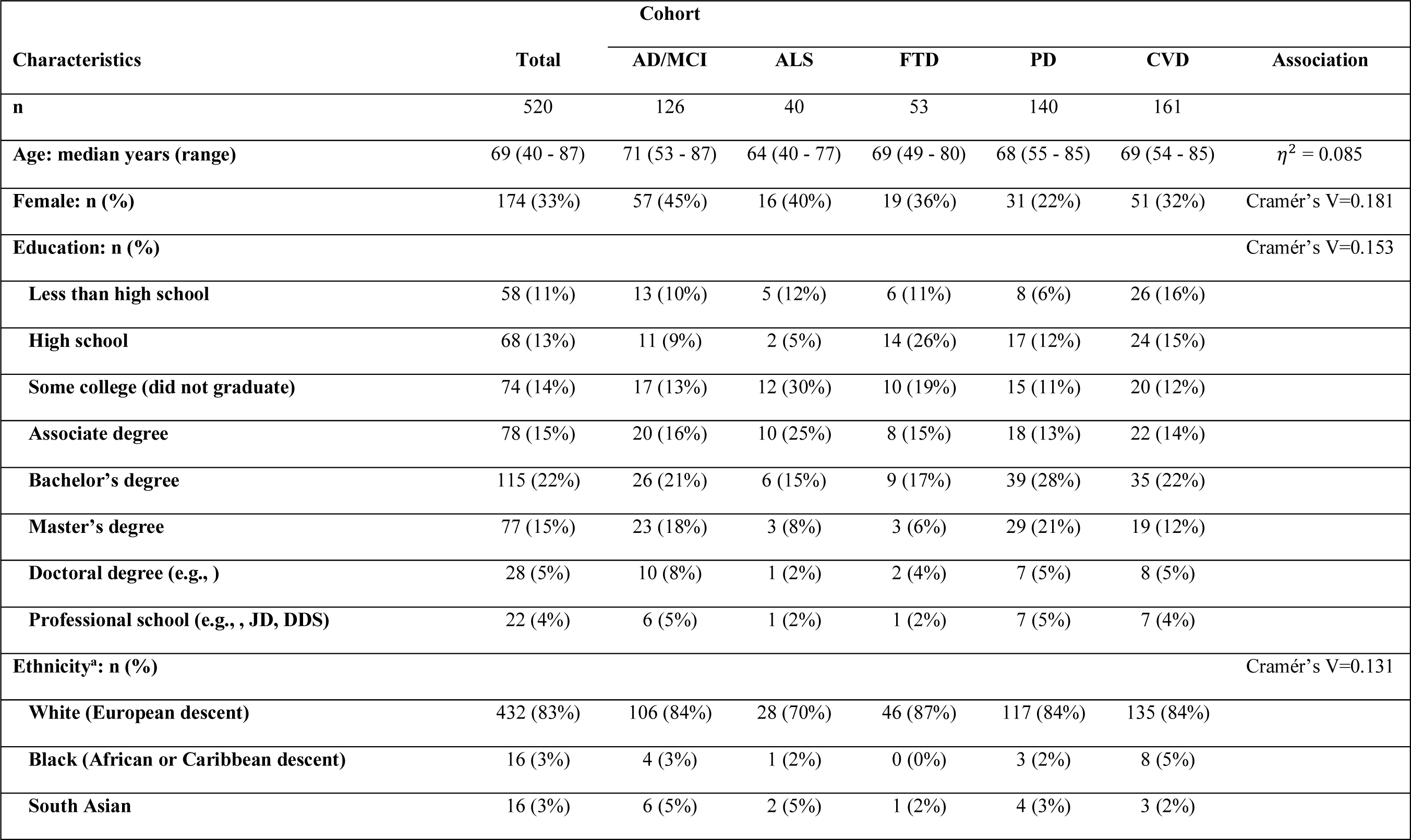
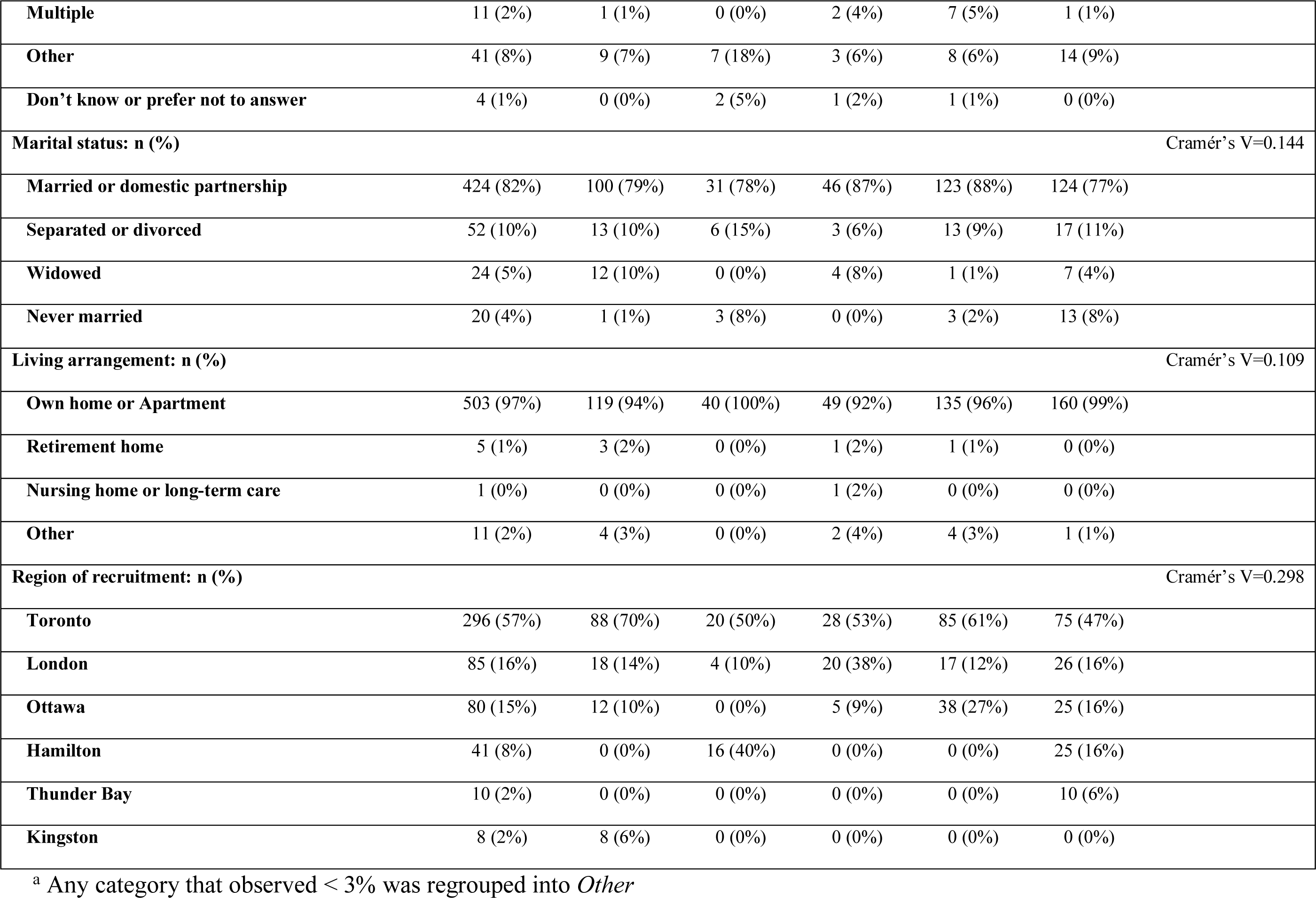
Participant demographics across the ONDRI sample and by disease cohort. AD/MCI = Alzheimer’s disease and amnestic mild cognitive impairment; ALS = amyotrophic lateral sclerosis; FTD = frontotemporal dementia spectrum disorder; PD = Parkinson’s disease; CVD = cerebrovascular disease.

**Table 3:** Study partner demographics and relation to participants across the ONDRI sample and by disease cohort. AD/MCI = Alzheimer’s disease and amnestic mild cognitive impairment; ALS = amyotrophic lateral sclerosis; FTD = frontotemporal dementia spectrum disorder; PD = Parkinson’s disease; CVD = cerebrovascular disease.

| Characteristics | Total | Cohort |  |  |  |  | Association |
| --- | --- | --- | --- | --- | --- | --- | --- |
|  |  | AD/MCI | ALS | FTD | PD | CVD |  |
| **n** | 520 | 126 | 40 | 53 | 140 | 161 |  |
| **Relationship to participant: n (%)** |  |  |  |  |  |  | Cramér's V=0.114 |
| Spouse or domestic partner | 398 (77%) | 88 (70%) | 30 (75%) | 41 (77%) | 118 (84%) | 121 (75%) |  |
| Child | 63 (12%) | 25 (20%) | 4 (10%) | 8 (15%) | 9 (6%) | 17 (11%) |  |
| Friend | 34 (7%) | 6 (5%) | 3 (8%) | 2 (4%) | 6 (4%) | 17 (11%) |  |
| Other relative | 25 (5%) | 7 (6%) | 3 (8%) | 2 (4%) | 7 (5%) | 6 (4%) |  |
| Age: median years (range) | 64 (19 - 87) | 66 (19 - 85) | 60 (26 - 77) | 62 (22 - 84) | 64 (22 - 85) | 65 (22 - 87) | $\eta^2 = 0.019$ |
| Female: n (%) | 389 (75%) | 86 (68%) | 26 (65%) | 41 (77%) | 115 (82%) | 121 (75%) | Cramér's V=0.132 |
| **Education: n (%)** |  |  |  |  |  |  | Cramér's V=0.158 |
| Less than high school | 43 (8%) | 10 (8%) | 5 (12%) | 4 (8%) | 6 (4%) | 18 (11%) |  |
| High school | 75 (14%) | 14 (11%) | 1 (2%) | 10 (19%) | 18 (13%) | 32 (20%) |  |
| Some college (did not graduate) | 57 (11%) | 14 (11%) | 5 (12%) | 8 (15%) | 15 (11%) | 15 (9%) |  |
| Associate degree | 92 (18%) | 16 (13%) | 13 (32%) | 13 (25%) | 26 (19%) | 24 (15%) |  |
| Bachelor's degree | 143 (28%) | 37 (29%) | 7 (18%) | 12 (23%) | 44 (31%) | 43 (27%) |  |
| Master's degree | 71 (14%) | 24 (19%) | 7 (18%) | 4 (8%) | 21 (15%) | 15 (9%) |  |
| Doctoral degree | 19 (4%) | 5 (4%) | 0 (0%) | 1 (2%) | 7 (5%) | 6 (4%) |  |
| **Professional school** | 19 (4%) | 6 (5%) | 1 (2%) | 1 (2%) | 3 (2%) | 8 (5%) |  |
| **Unknown** | 1 (0%) | 0 (0%) | 1 (2%) | 0 (0%) | 0 (0%) | 0 (0%) |  |
| **Lives with participant: n (%)** | 419 (81%) | 96 (76%) | 33 (82%) | 41 (77%) | 122 (87%) | 127 (79%) | Cramér's V=0.109 |
| **Time living with participant (if live together = yes): median years (range)** | 36 (<1 - 65) | 35 (<1 - 65) | 30 (1 - 55) | 40 (4 - 57) | 36 (<1 - 63) | 38 (1 - 63) | $\eta^2 = 0.021$ |
| **Time with participant per week (if live together = no): median hours (range)** | 6 (0.5 - 112) | 8 (1 - 112) | 15 (3 - 80) | 4.5 (1 - 16) | 3.5 (0.5 - 23) | 7.5 (1 - 108) | $\eta^2 = 0.066$ |

Overall, 67% of ONDRI participants were male, 83% identified their ethnicity as white, 76% were married, and 97% lived in their own homes. As for the study partners, 77% were in spousal or domestic relationships with the participant, 75% were female, and 80% lived with the participant. Age varied moderately between diseases (*η*2 = 0.085), and measures of association of categorical variables with disease varied, with the largest association between cohort and region of recruitment (Cramér’s V = 0.298).

### Clinical characteristics

Table 4 summarizes general and disease-specific measures for participants in each of the disease cohorts. Variation between the five cohorts (collapsing across subtypes) was large for MoCA total score (*η*^2^ = 0.219). Just over half of participants in both the CVD and PD cohorts scored 26 or higher on the MoCA (53% and 58%, respectively).

**Table 4:**
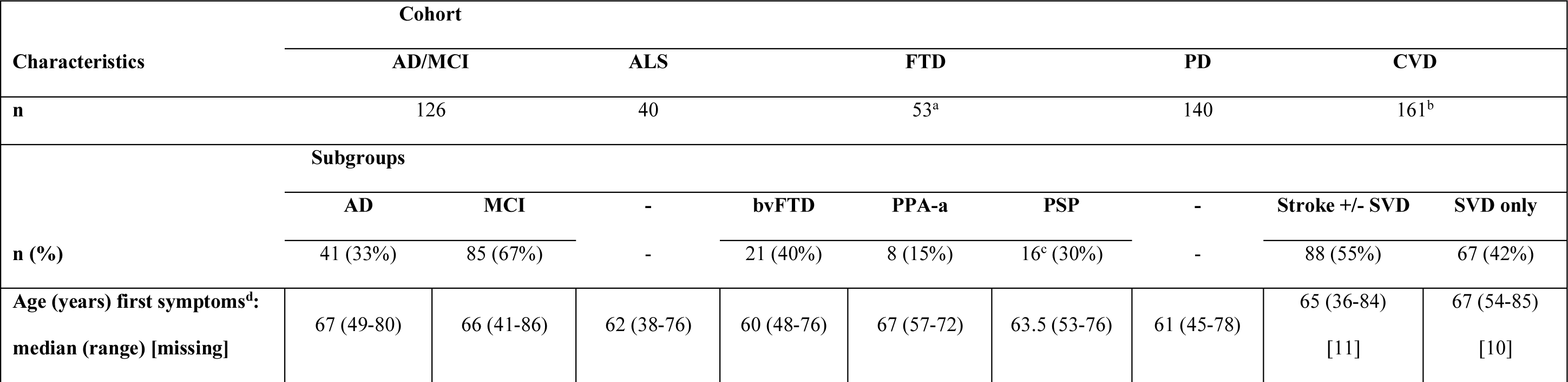
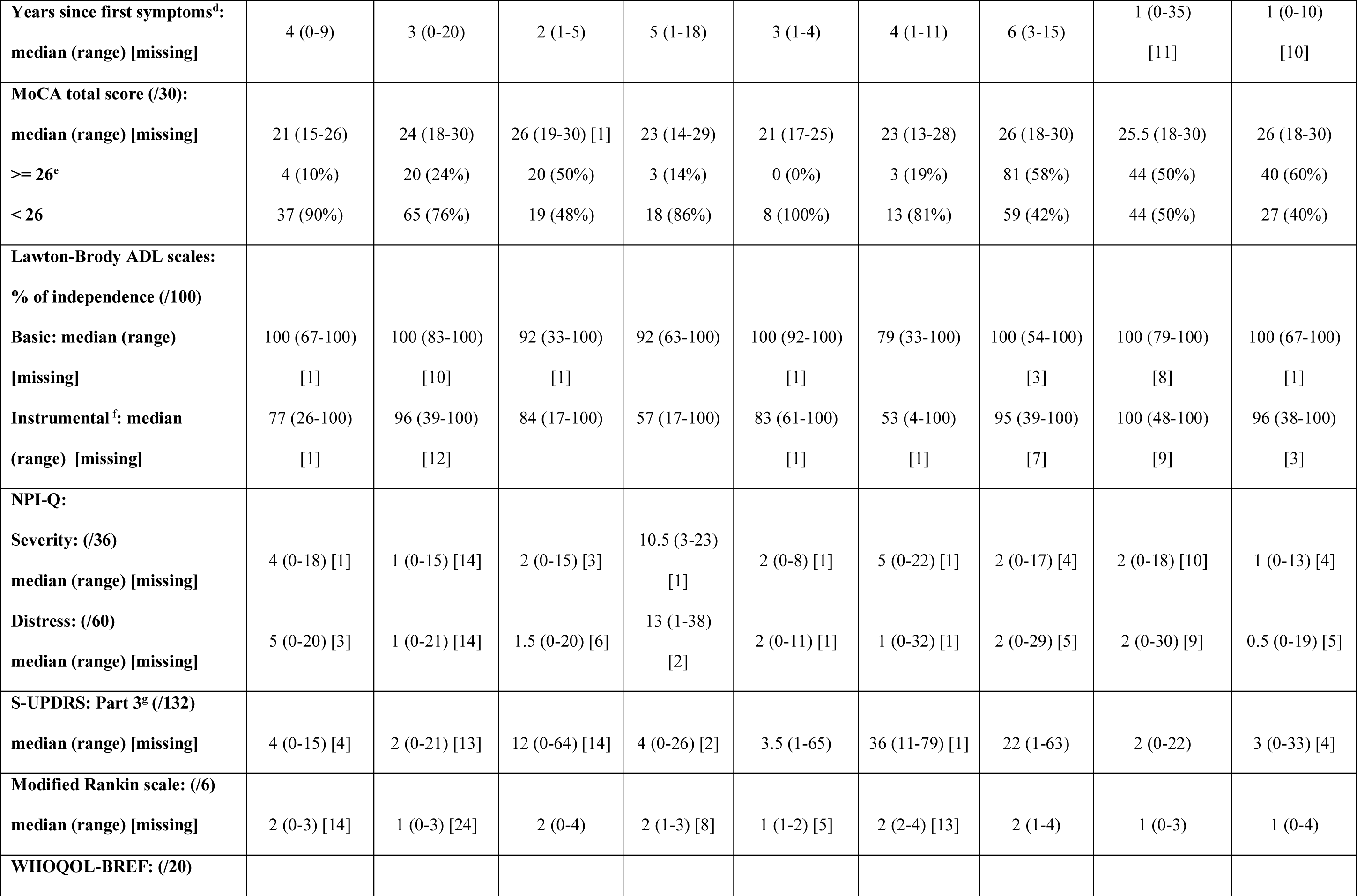
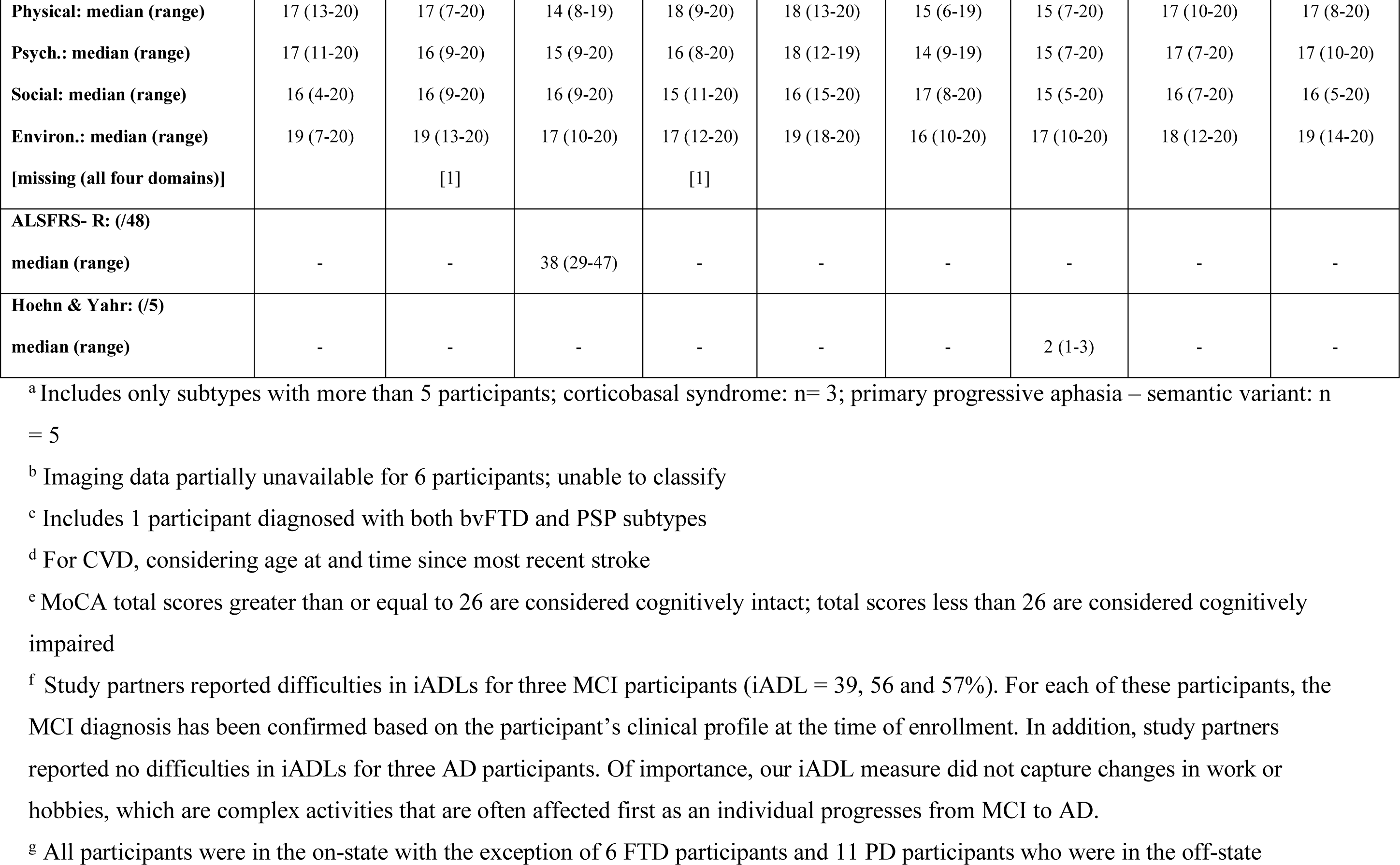
Participant disease-specific characteristics, by disease cohorts and subtypes, where applicable. AD = Alzheimer’s disease; MCI = amnestic mild cognitive impairment; ALS = amyotrophic lateral sclerosis; FTD = frontotemporal dementia spectrum disorder; bvFTD = behavioural variant FTD; PPA-a = progressive primary aphasia – agrammatic variant; PSP = progressive supranuclear palsy; PD = Parkinson’s disease; CVD = cerebrovascular disease; SVD = small vessel disease; MoCA = Montreal Cognitive Assessment; ADL = Activities of Daily Living; NPI-Q = Neuropsychiatric Inventory Questionnaire; S-UPDRS = Movement Disorder Society revised Unified Parkinson’s Disease Rating Scale; WHOQOL-BREF = Abbreviated World Health Organization Quality of Life assessment; Psych. = Psychological; Environ. = Environmental; ALSFRS-R = ALS Functional Rating Scale – Revised. A higher score on the MoCA, Lawton-Brody ADL scales, WHOQOL-BREF, and ALSFRS-R indicates a closer resemblance of normalcy. A higher score on the NPI-Q, S-UPDRS, modified Rankin scale and Hoehn & Yahr indicates a higher severity of measured symptoms.

### Multivariate relationships between demographic, study partner, and MoCA variables

In order to understand dependencies among clinical and demographic variables, which is required for modelling their role as factors in cognitive and motor impairments, we identified multivariate associations using MCA.

Two components explained 25.8% of the total variance (17.2% and 8.7% separately). As illustrated in Figure 3(A), Component 1 indicates that participants who are not married, have non-spousal study partners (i.e., friends, adult children, and other relatives), and do not live with their study partner are associated. Component 2 shows that male participants, female study partners, younger participants, younger study partners, higher MoCA total scores, and higher education are associated. Black, south Asian, and other ethnicities are associated with both components.

**Figure 3:**
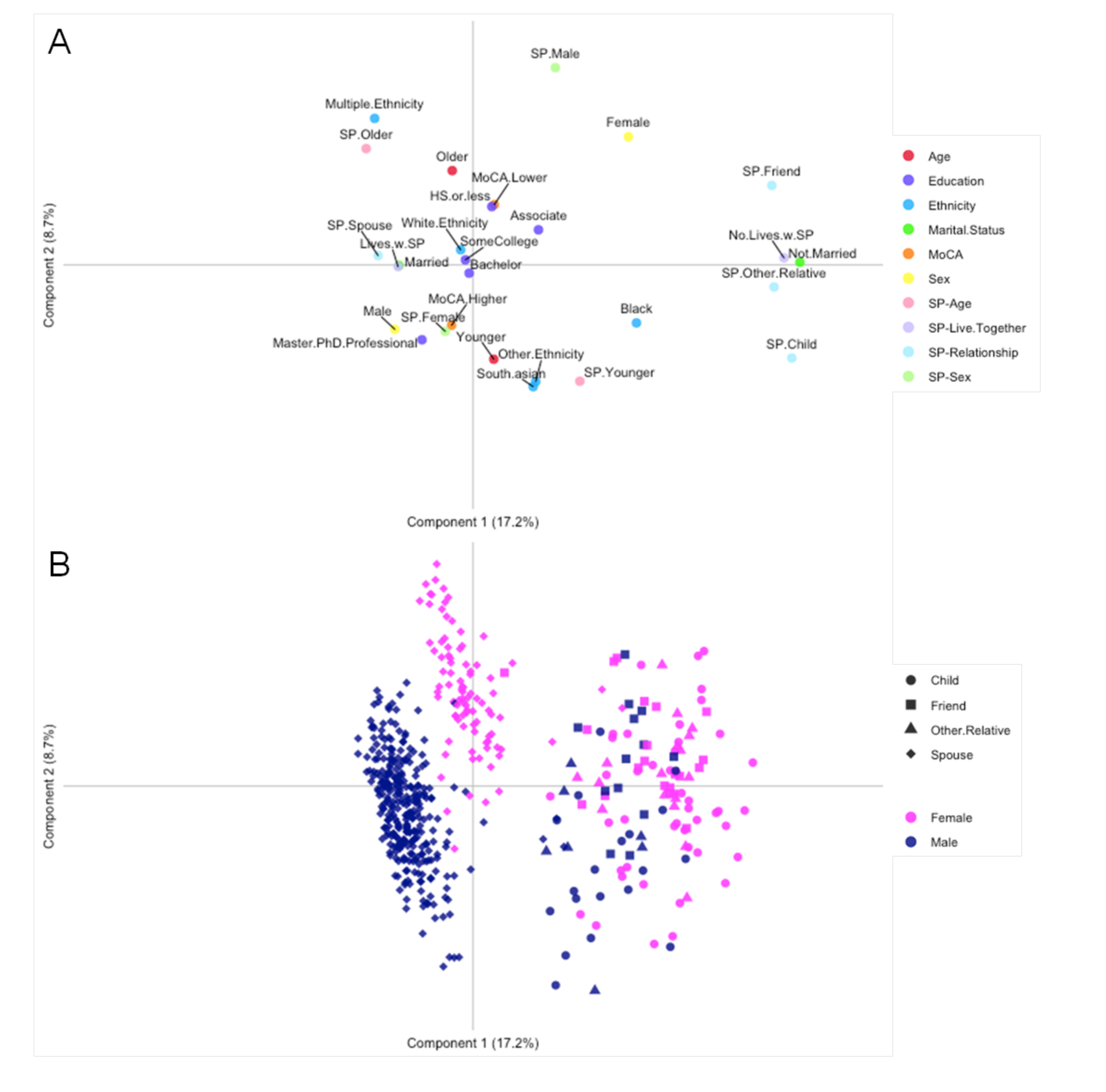
Component maps of Multiple Component Analysis results. A) Variable component scores; B) Participant component scores. Response categories with fewer observed responses have been combined: For education, ‘HS.or.less’ includes participants with less than a high school education, as well as those who earned their diploma; ‘Master..Professional’ includes participants in any of the ‘Master’s degree’, ‘Doctoral degree’, or ‘Professional school’ response categories. For ethnicity, ‘Other’ includes response categories ‘don’t know’ or ‘prefer not to answer.’ For marital status, ‘Not.Married’ includes participants who were separated, divorced, widowed, or never married. MoCA = Montreal Cognitive Assessment total score; SP = study partner; HS = high school; AD/MCI = Alzheimer’s disease and amnestic mild cognitive impairment; ALS = amyotrophic lateral sclerosis; FTD = frontotemporal dementia spectrum disorder; PD = Parkinson’s disease; CVD = cerebrovascular disease.

Reflecting these associations, participants clustered into three groups, as illustrated in Figure 3(B). The right hand cluster included 127 participants (24%) who were not married, had a non-spousal study partner, or did not live with their study partner. Most of this group was female (67%) and 24% were of black, south Asian, or other ethnicity. Of the remaining 393 participants (76% of all 520), 386 (98%) were married, had a spousal study partner, and lived with their study partner; 46 (12%) were of black, south Asian, or other ethnicity. The upper left cluster included 89 participants (17% of all 520) of whom 86 (97%) were female and 33 (37%) had attained a Bachelor’s degree or higher, while the lower left cluster included 304 participants (58% of all 520) of whom 3 (1%) were female and 159 (52%) had attained a Bachelor’s degree or higher.

### Participants missing platform data

In total, 436 participants (84%) provided usable data for all platforms except retinal imaging. In addition to pre-specified exceptions, the retinal imaging platform encountered service agreement delays with the Toronto recruitment sites, delaying the start of data collection. As a direct result, 156 participants (30%) did not complete the retinal imaging assessment at baseline. Including retinal imaging, 231 participants (44%) provided usable data across all assessment platforms.

### Data collection time window

Overall, 430 participants (83%) completed baseline visits within the targeted eight weeks after providing consent, with a median time frame of 6.14 weeks. Most cases in which the data collection was outside the eight-week window were as a result of scheduling conflicts. Two data collection decisions also had an impact: (1) MRI scans for the neuroimaging assessment that did not meet quality control standards for processing were re-acquired, and 8 of the 10 re- acquisitions (80%) were past the eight week window, and (2) because of the aforementioned retinal imaging service agreement delays, participants recruited within the six months before the agreements were in place were asked to complete the retinal imaging assessment at that time, which accounts for 14 of the 58 assessments (24%) beyond the eight week time window.

## Discussion

ONDRI collected a wide array of data across multiple assessment platforms with which researchers can study clinical and biological features within, between, and across multiple common neurodegenerative diseases associated with aging and cerebrovascular disease. The distribution of age, sex, education, and ethnicity are similar to other studies of specific neurodegenerative diseases^2-5^.

The MCA highlighted that requiring a study partner may have influenced sample demographic characteristics. Consistent with other studies including a study partner^40^, a majority of ONDRI participants had a spousal study partner. Work obligations and logistical challenges (e.g., not living with the participant) often interfere with the ability of other relatives or friends to participate^40^. Participants with spousal study partners are more often male, white, and more educated^40^, as was reflected in our sample. Further, previous research has shown that individuals who identify as non-white are more likely to support non-spousal family members or friends^40^. The MCA also identified associations between age, education, and MoCA total score. Consistent with previous research on the MoCA^41^, participants with higher MoCA scores were generally younger and had attained higher levels of education. Studies of cognition must consider the interplay between assessment measures and potentially influential demographic characteristics in order to draw appropriately nuanced conclusions regarding impairment, preservation, or decline. Some disease cohorts were recruited more heavily from particular geographic regions of Ontario. For example, ALS participants were more heavily recruited from Hamilton, a city known for its large share of the manufacturing industry, than other regions. A higher proportion of technical degrees in this cohort may be attributed to this recruitment decision and, importantly, level of education and occupational attainment may influence performance on cognitive assessments^42^. As a result, differences between cohorts may also be partially attributable to recruitment patterns across regions with different demographic distributions.

ONDRI enrolled participants across a range of disease severities, though participants in earlier stages of disease, with features that suggest an increased likelihood of participation in comprehensive follow-up, predominate each cohort. Specifically, in addition to the MCI subgroup, more than half of participants in the AD subgroup had reportedly intact basic activities of daily living and relatively mild deficiencies noted on instrumental activities of daily living, showing early stages of impairment in this cohort. The majority of ALS participants were diagnosed with limb onset ALS and therefore had better prognosis than those with bulbar onset, depending on other factors such as age^43^. The majority of FTD participants had no more than slight symptoms on non-motor experiences of daily living. Over half of participants in the PD cohort had no more than slight symptoms on most aspects of daily living, indicative of an opportunity to observe disease progression over follow-up visits^44^. Nearly three-quarters of participants in the CVD cohort scored 0 or 1 on the modified Rankin scale, suggesting few symptoms or activity of daily living restrictions three or more months post-stroke, and increasing the likelihood of their participation across all data collection timepoints^45^.

Heterogeneity within disease was designed in order to capture a wide spectrum of clinical presentations: (1) both typical and atypical presentations of Alzheimer’s disease, as well as amnestic mild cognitive impairment are represented in the AD/MCI cohort, (2) the FTD cohort included five disease subtypes including behavioural variant, agrammatic/non-fluent variant of primary progressive aphasia, and progressive supranuclear palsy as the most frequent, (3) the CVD and PD cohorts enrolled both cognitively intact and impaired participants. Minor variances in clinical characteristics were also induced through protocol amendments. Lastly, unintended features emerged during initial data explorations including incidental strokes observed in a few ALS, FTD, and PD participants. However, a research neuroradiologist confirmed that these incidental strokes did not interfere with disease-specific symptoms and the participants remained in the cohorts to which they were recruited.

Two approaches to this ONDRI sample that capitalize on this diversity are indicated. For analyses within disease cohorts, explicit acknowledgement of heterogeneity through investigation of subgroups could illuminate important differences within standard clinical diagnoses. Alternatively, a disease-agnostic approach that considers ONDRI participants as a sample of individuals with neurodegenerative or cerebrovascular disease by removing disease labels may identify some features that align well with baseline diagnosis, and other patterns that are more suggestive of mixed disease. Mixed disease is being recognized as the norm rather than the exception with respect to its contribution to cognitive decline in individuals over age 65 through several longitudinal studies of aging and dementia^6^.

Studies of vascular cognitive impairment often recruit from memory clinics leading to samples with cognitive impairment at baseline. However, most participants in the CVD cohort were recruited from stroke clinics and include individuals with and without memory concerns. The inclusion of eight participants recruited for the AD/MCI cohort that were subsequently enrolled into the CVD cohort upon discovery of vascular pathologies at baseline are the exception. Analysis of this cohort may reveal implications of the different routes of entry between these eight participants and those recruited through stroke clinics.

Enrollment strategies and criteria played an important role in defining the sample, most notably for the AD/MCI cohort. Disease-specific requirements and a minimum MoCA threshold together were responsible for 56% of potential participants being excluded from this cohort. We propose two possible reasons for this phenomenon.

First, ONDRI recruited individuals through clinicians at tertiary clinics. However, when memory clinic training programs were introduced in Canada, primary-care clinics began providing effective early services for individuals with straightforward cognitive impairment^46^. While this approach increased capacity for care of individuals with cognitive decline, it also created a shift in patient characteristics and clinical presentation in tertiary clinics. As a result, fewer individuals with typical presentation early in the disease course are available for recruitment from tertiary clinics. While treatment for some neurodegenerative diseases such as PD remains at the tertiary level^47^, future studies of Alzheimer’s disease and/or mild cognitive impairment should consider recruitment from memory clinics at the primary-care level.

Second, standard criteria for the diagnosis of Alzheimer’s disease or mild cognitive impairment remain fundamentally clinical in Canada and best practice guidelines do not require MRI^48^. As a result, multiple individuals recruited to the AD/MCI cohort presented other possible causes for cognitive decline on MRI, including vascular pathologies and markers consistent with traumatic brain injury. Such findings excluded 15 participants from the ONDRI AD/MCI cohort^21^ and reinforced the importance of neuroimaging for both research studies and in order for patients to receive more appropriate care.

Apart from adapted collection for retinal imaging, more than 80% of participants completed all assessments within eight weeks. This feat, which required stamina and dedication from participants and their study partners, indicates that the protocol was manageable for individuals in early stage neurodegenerative or cerebrovascular disease. Future research using ONDRI’s longitudinal data will be able to explore whether disease progression is related to attrition and at what stage a protocol such as this becomes too burdensome. It will also facilitate the identification of contributing factors to both slow and rapid disease progressors within and across the different neurodegenerative diseases, which is an important confound to control for in future clinical trials of potential disease-modifying agents.

Some participants are missing data for an entire assessment platform, while other missing data are intermittent throughout a platform’s measures. Participants can be missing data for various reasons, however missing data as a result of cognitive, behavioural, or physical impairment are of prominent interest as these may be more frequent among participants at more advanced stages of disease. As a result, complete case analysis may be misleading as estimates of parameters of interest would tend to indicate associations relevant only to participants with better performance. Informative missing data codes are embedded in all ONDRI data sets to assist researchers in determining appropriate approaches for accommodating missing values that maintain generalizable conclusions.

ONDRI data underwent extensive cleaning and quality control pipelines to identify and correct data entry or processing errors^26–31^, or document anomalies^34^, thereby increasing the accuracy and integrity of data and consequently reducing the risk of incorrect conclusions. While such protocols can be time consuming, they serve a two-fold benefit: more accurate results due to correction of erroneously recorded data and more nuanced analyses sensitive to idiosyncrasies in the sample.

Translational neurodegenerative disease research recognizes the importance of multi-disciplinary team science, as shown through multiple prior initiatives^2–5^. Large scale and broadly accessible data encourages additional analyses on curated data but also allows researchers to merge data from multiple studies, leverage a constellation of initiatives, and accelerate discovery through broader integrations of data and wider collaborations of researchers^49^.

Most neurodegeneration initiatives consider a particular cohort or disease, and because independent studies deploy different measures, assessments, and methodological specifications, cross-disease comparisons between studies are limited^50^. ONDRI is in a distinct position to overcome these challenges because of its breadth of neurodegenerative diseases sampled and its depth of harmonized phenotypic characterization. In addition, data from the biomarker and pathology platform are being ascertained in order to characterize participants based on their underlying proteinopathies (e.g., antemortem plasma P181-Tau, post-mortem immunohistochemical neuropathological diagnosis), degree of neurodegeneration (e.g., plasma NfL), and markers of neuroinflammation (e.g., plasma GFAP). This data will also expand on the capacity to identify individuals with pure vs. mixed contributions to cognitive and motor decline. ONDRI data from 520 participants with neurodegenerative or cerebrovascular disease broadly captured multiple components comprising the disease profile. The outcome of the executed protocol and baseline demographic and select clinical characteristics of the participants are described to promote deep investigation into many elements of clinical, pathological, and societal mechanisms. These data will be available to the scientific community through the Ontario Brain Institute later this year.

## Data Availability

All ONDRI data, including but not limited to data used in this article, will be available to the scientific community through the Ontario Brain Institute later this year. Please see the Ontario Brain Institute website for more information.

https://braininstitute.ca/

## Acknowledgements

This work was completed on behalf of the Ontario Neurodegenerative Disease Research Initiative (ONDRI). The authors would like to express their sincere gratitude to the participants, their study partners, and their families for the time and effort given to this study.

The authors and investigators would like to acknowledge ONDRI’s project management team, past and present: Susan Boyd, Lisa Desalaiz, Catarina Downey, Stephanie Hetherington, Heather Hink, Ruth Kruger, Sean Lucas, Donna McBain, Andrea Richter, Michelle Vanderspank, Joanna White, and Ashley Wilcox, and the coordinators at each site. The authors would also like to acknowledge the recruitment sites in Ontario, Canada: Baycrest Health Sciences, Toronto; Centre for Addiction and Mental Health, Toronto; Elizabeth Bruyère Hospital, Ottawa; Hamilton General Hospital, Hamilton; Hotel Dieu Hospital, Kingston; London Health Sciences Centre, London; McMaster Medical Centre, Hamilton; Parkwood Institute, London; Providence Care

Hospital, Kingston; St. Michael’s Hospital, Toronto; Sunnybrook Health Sciences Centre, Toronto; The Ottawa Hospital, Ottawa; Thunder Bay Regional Health Sciences Centre, Thunder Bay; and Toronto Western Hospital (University Health Network), Toronto.

This research was conducted with the support of the Ontario Brain Institute, an independent non-profit corporation, funded partially by the Ontario Government. The opinions, results, and conclusions are those of the authors and no endorsement by the Ontario Brain Institute is intended or should be inferred.

Matching funds were provided by participating hospital and research institute foundations, including the Baycrest Foundation, Bruyère Research Institute, Centre for Addiction and Mental Health Foundation, London Health Sciences Foundation, McMaster University Faculty of Health Sciences, Ottawa Brain and Mind Research Institute, Queen’s University Faculty of Health Sciences, Providence Care (Kingston), St. Michael’s Hospital, Sunnybrook Health Sciences Foundation, the Thunder Bay Regional Health Sciences Centre, the University of Ottawa Faculty of Medicine, and the Windsor/Essex County ALS Association. Also, the Temerty Family Foundation provided the major infrastructure matching funds.

In addition, CF is supported by Vielight Inc, Hoffman LaRoche, Brain Canada, PCORI and the Heather and Eric Donnelly Endowment; MJ is supported by CIHR, Parkinson Society Canada, MITACS, OCE, Research Council of Norway, Abbvie, Allergan, Merz Pharma; SK is supported by Brain and Behavior Foundation, National institute on Ageing, BrightFocus Foundation, Brain Canada, Canadian Institute of Health Research, Centre for Ageing and Brain Health Innovation, Weston Brain Institute, Centre for Mental Health and Addiction Foundation and University of Toronto; DSP is supported by CIHR; ACR is supported by Canadian Institutes of Health Research Fellowship, Parkinson Society Canada, (NIH) National Institute on Deafness and Other Communication Disorders (1R21DC017255-01).

